# Machine Learning Model for Predicting Outcomes of Biologic Therapy in Psoriasis

**DOI:** 10.1101/2021.12.05.21267219

**Authors:** Amy X. Du, Zarqa Ali, Kawa K. Ajgeiy, Maiken G. Dalager, Tomas N. Dam, Alexander Egebjerg, Christoffer V. S. Nissen, Lone Skov, Simon Francis Thomsen, Sepideh Emam, Robert Gniadecki

## Abstract

**Background:** Biological agents used for the therapy of psoriasis lose efficacy over time, which leads to discontinuation of the drug. Optimization of long-term biologic treatment is an area of medical need but there are currently no prediction tools for biologic drug discontinuation.

**Objective:** To compare the accuracy of the risk factor-based frequentist statistical model to machine learning to predict the 5-year probability of biologic drug discontinuation.

**Methods:** The national Danish psoriasis biologic therapy registry, Dermbio, comprising 6,172 treatment series with anti-TNF (Etanercept, Infliximab, Adalimumab), Ustekinumab, Guselkumab and anti-IL17 (Secukinumab and Ixekizumab) in 3,388 unique patients was used as data source. Hazard ratios (HR) were computed for all available predictive factors using Cox regression analysis. Different machine learning (ML) models for the prediction of 5-year risk of drug discontinuation were trained using the 5-fold cross validation technique and using 10 clinical features routinely assessed in psoriasis patients as input variables. Model performance was assessed using the area under the receiver operating characteristic curve (AUC).

**Results:** The lowest 5-year risk of discontinuation was associated with therapy with ustekinumab or ixekizumab, male sex and no previous exposure to biologic therapy. The predictive model based on those risk factors had an AUC of 0.61. The best ML model (gradient boosted tree) had an AUC of 0.85.

**Conclusions:** A machine learning-based approach, more than a statistical model, accurately predicts the risk of discontinuation of biologic therapy based on simple patient variables available in clinical practice. ML might be incorporated into clinical decision making.

## Introduction

The widespread implementation of biologic therapy in psoriasis has changed the landscape of treatment for this disease. Prior to their use, the journey to remission was lengthy, required multiple trials of topical and systemic agents, and posed a significant risk of drug toxicity to patients living with psoriasis. Presently, biologic therapies (comprising tumour necrosis factor (TNF) inhibitors, interleukin (IL)-17A inhibitors, IL-23 or IL12/23 inhibitors) have been shown to have PASI 90 responses in up to 80% of patients, and PASI 75 responses in up to 90% of patients.^1,2^

Large-scale registry studies have shown that, although the short-term efficacy of biologics is comparable to what was shown in phase III clinical trials, long-term therapy is associated with the gradual decrease of efficacy, leading to discontinuation of treatment in a significant proportion of psoriasis patients.^3–5^ In a meta-analysis of the registry data looking at the risk of discontinuation, discontinuation may be as high as 50-65% over 5 years, depending on the drug.^6^ Therefore, the current clinical challenge is to optimize strategies to predict long-term efficacy of biologics.

Despite an extensive amount of data describing the efficacy of biologics, the therapeutic decision-making process is still largely based on trial-and-error. Several risk factors of biologic discontinuation have been identified (e.g., obesity, female sex, biologic class),^5,7–11^ but they have not been incorporated into clinical practice because of their low predictive value. A precision medicine approach, where patients’ unique characteristics are accounted for, would allow for better assessments and predictions to determine the most appropriate drug. Such an approach would assist physicians in selecting a biologic with the highest chance of continuation for their patient right from the initiation of treatment, thereby saving time and resources for both the patient and healthcare system.

Machine learning (ML) is an emerging tool that can detect hidden complex patterns and has been used with success to predict future trajectories of patients’ health in diverse areas of healthcare.^12–17^ ML techniques are increasingly being approved by the regulatory agencies to assist decision making in medicine.^18^ Our group has explored ML for prediction of therapy outcomes in psoriasis, and we found that ML predicted drug discontinuation risk with 82% accuracy.^19^ However, this study was preliminary and based on a limited dataset.

In our current study, we compared the accuracy of prediction of biologic drug discontinuation rate between the classic statistical risk factor modelling to different ML algorithms. By analyzing data from the Danish psoriasis registry, Dermbio, we show that ML was superior to a classical statistical risk factor model at predicting therapy outcome with a high accuracy.

## Methods

### Data Source and Study Population

Individual patient data were obtained from Dermbio, a longitudinal registry of patients with moderate-to-severe psoriasis, who received biologic treatment in Denmark from 2003 to present. The structure of and the data available in the Dermbio registry have been previously described in detail.^5,8,20^ Briefly, the database was created to monitor the safety and efficacy of biologic drugs used for psoriasis treatment in Denmark, and since 2007, nationwide registration by dermatologists has been mandatory upon the initiation or change of any biologic therapy.

The registry includes data in the form of treatment series, delineating the name of biologic initiated/changed and for what length of time, reason for discontinuation, disease activity (Psoriasis Area Severity Index (PASI) Dermatology Life Quality Index (DLQI)), patient demographic data (age, sex, weight, height, body mass index (BMI)), concomitant methotrexate therapy, presence of psoriatic arthritis and other comorbidities.^20^ We included data on patients treated with adalimumab, etanercept, guselkumab, infliximab, ixekizumab, secukinumab, or ustekinumab. The cut-off date for treatment series for our study was October 1, 2020.

### Data Pre-Processing

To ensure accurate analyses, the following steps were taken in the data preparation process: (1) Correction of data formats to ensure consistency and labeling missing data as such, rather than by default “zero” values; (2) Variables with a large number of missing values were removed, e.g., blood pressure, smoking status; (3) Erroneous data, e.g., treatment series that had a start date after the end date or duplicate series, were removed; (4) Treatment series of duration less than one month were removed; (5) Data from two consecutive series were merged for the same biologic if the period between the cycles was less than one month.

As biosimilars have become available for Enbrel® (Benepali®) and Remicade® (Inflectra®, Remsima®, Zessly®), and because previous studies did not demonstrate any differences in outcomes between originators and biosimilars^5^, we did not differentiate between the biosimilar and originator data, labeling them both by the generic name of the biologic, i.e. etanercept and infliximab, respectively. If a patient was transitioned off the originator to the biosimilar, the data were merged into one treatment series.

### Descriptive Statistical Analyses

We present patient characteristics as means with standard deviations for continuous variables, and frequencies with percentages for categorical variables. We also include percentages delineating the completeness of data for each variable, as more data was captured for some variables than others. If more than one value was available for the same time point, as in the case with patient weight, the values were averaged. For baseline PASI and DLQI scores, the value at the patient’s first registered treatment series was included.

Drug survival analyses were performed using Cox regression, reporting hazard ratio (HR) and 95% confidence intervals (CI), as described previously.^21^ We considered a statistically significant P-value to be < 0.05. Based on Cox regression data a predictive nomogram was created to estimate the probability of remaining on the drug at 1-year, 3-years, and 5-years.

All statistical analyses were performed, and figures were created, using the following packages in RStudio 4.0.5 (Boston, MA, USA): dplyr (v1.0.5; Wickham et al., 2021), ggplot2 (v3.3.3; Wickham et al., 2020), rms (v6.2.0; Harrell, 2021), survival (v3.2.10; Therneau et al., 2021), survminer (v0.4.9; Kassambara et al., 2021).

### Machine Learning Models

Supervised machine learning techniques were used to extract a stochastic model from the patients’ dataset to predict outcomes of biologic therapy (risk of discontinuation). Based on our preliminary modeling data^19^ we considered six different modelling techniques: (1) Generalized Linear Model (GLM), (2) Naive Bayes, (3) Deep Learning, (4) Decision Tree (DT), (5) Random Forest, and (6) Gradient Boosted Trees.

The input data comprised sex, age, weight, height, concomitant methotrexate therapy, PASI score, DLQI score, presence of psoriatic arthritis (PsA), type of biologic therapy, previous therapies. In order to assess the effectiveness of the models, we applied the 5-fold cross validation technique as part of the performance analysis procedure. This technique randomly partitions a set of examples into five, non-overlapping, sets. Subsequently, a model is inferred using each set over five iterations and the remaining set is used to evaluate the model based upon the accuracy measure. This is defined in the next section. Since each iteration uses a different set for the evaluation, the final accuracy score is equal to the mean of the five accuracy scores. This assists in evaluating the performance of our machine learning approach on some unseen data and identifies overfitting or underfitting related issues.

ML modeling was done using the glm2 package in R, GLM and DT packages in Python, and RapidMiner simulator.

### Performance Analysis

In order to statistically evaluate the performance of learning approaches used in this study, we produced confusion tables as a result of classification procedure containing True Positives (TP), False Negatives (FN), True Negatives (TN) and False Positives (FP), which were used to calculate diagnostic accuracy with the formula (TP+TN)/(TP+FP+FN+TN).

Receiver operating characteristic (ROC) curves were constructed using R libraries multiROC and ggplot2 (downloaded from https://cran.r-project.org/web/packages/) to visualize diagnostic utility of the machine learning algorithms. Area under the curve (AUC) was calculated to determine which model provided the best prediction by measuring how true positive rate (recall) and false positive rate trade off. Of note, the ROC-AUCs were calculated by keeping a class and stacking the rest of the groups together, thus converting the multi-class classification into binary classification^.22^

## Results

### Patient Characteristics

A total of 3,388 unique patients were included in our study, most of whom received adalimumab (n = 1871) followed by ustekinumab (n = 1499) and secukinumab (n = 841) **(Table 1)**. Among those, 57.2% of patients received only one biologic therapy, whereas the others were treated with two (21.8%) or three or more biologics (21.0%).

**Table 1.** Patient characteristics of each biologic and total group. Completeness of data for each variable is indicated in italics.

| Characteristic | Adalimumab<br>(n = 1871) | Etanercept<br>(n = 815) | Guselkumab<br>(n = 87) | Infliximab<br>(n = 389) | Ixekizumab<br>(n = 206) | Secukinumab<br>(n = 841) | Ustekinumab<br>(n = 1499) | All Patients<br>(n = 3388) |
| --- | --- | --- | --- | --- | --- | --- | --- | --- |
| Sex, n (%) | Female: 713 (38.2)<br>Male: 1154 (61.8)<br>*99.8% complete* | Female: 310 (38.0)<br>Male: 505 (62.0)<br>*100% complete* | Female: 40 (46.0)<br>Male: 47 (54.0)<br>*100% complete* | Female: 158 (40.6)<br>Male: 231 (59.4)<br>*100% complete* | Female: 80 (38.3)<br>Male: 126 (61.7)<br>*100% complete* | Female: 345 (41.0)<br>Male: 496 (59.0)<br>*100% complete* | Female: 590 (39.4)<br>Male: 909 (60.6)<br>*100% complete* | Female: 1261 (37.3)<br>Male: 2124 (62.7)<br>*99.9% complete* |
| Mean age at treatment series, years (SD) | 43.0 (15.1)<br>*100% complete* | 46.3 (14.7)<br>*100% complete* | 47.1 (13.8)<br>*100% complete* | 46.6 (13.7)<br>*100% complete* | 48.7 (14.0)<br>*100% complete* | 46.5 (14.4)<br>*100% complete* | 44.6 (15.1)<br>*100% complete* | 44.8 (14.9)<br>*100% complete* |
| Mean age at diagnosis, years (SD) | 25.5 (13.9)<br>*69.5% complete* | 27.0 (14.9)<br>*80.7% complete* | 24.3 (14.0)<br>*64.4% complete* | 27.0 (15.1)<br>*85.3% complete* | 25.4 (13.1)<br>*64.1% complete* | 25.0 (13.5)<br>*68.7% complete* | 24.4 (14.0)<br>*65.6% complete* | 25.4 (14.2)<br>*71.8% complete* |
| Mean age at first biologic, years (SD) | 43.1 (14.6)<br>*100% complete* | 45.2 (14.9)<br>*100% complete* | 42.4 (13.4)<br>*100% complete* | 44.9 (14.4)<br>*100% complete* | 44.9 (13.9)<br>*100% complete* | 43.8 (14.2)<br>*100% complete* | 43.1 (15.1)<br>*100% complete* | 43.9 (14.8)<br>*100% complete* |
| Mean weight, kg (SD) | 92.7 (70.8)<br>*89.8% complete* | 88.9 (22.4)<br>*87.7% complete* | 98.8 (26.7)<br>*74.7% complete* | 95.0 (26.3)<br>*92.8% complete* | 96.6 (25.5)<br>*81.1% complete* | 92.9 (43.8)<br>*91.2% complete* | 91.0 (33.7)<br>*91.5% complete* | 91.6 (57.4)<br>*88.9% complete* |
| Mean BMI, kg/m<sup>2</sup> (SD) | 28.5 (13.7)<br>*81.7% complete* | 26.3 (12.2)<br>*73.9% complete* | 29.0 (12.0)<br>*72.4% complete* | 30.5 (9.7)<br>*81.0% complete* | 28.3 (11.2)<br>*74.8% complete* | 29.6 (17.6)<br>*80.4% complete* | 29.1 (12.7)<br>*80.3% complete* | 28.5 (13.2)<br>*76.7% complete* |
| Concurrent PsA, n (%) | Yes: 688 (38.7)<br>No: 1091 (61.3)<br>*95.1% complete* | Yes: 368 (45.2)<br>No: 447 (54.8)<br>*100% complete* | Yes: 37 (44.0)<br>No: 47 (56.0)<br>*96.6% complete* | Yes: 186 (48.7)<br>No: 196 (51.3)<br>*98.2% complete* | Yes: 78 (39.4)<br>No: 120 (60.6)<br>*96.1% complete* | Yes: 356 (45.8)<br>No: 398 (54.2)<br>*89.7% complete* | Yes: 340 (23.5)<br>No: 1109 (76.5)<br>*96.7% complete* | Yes: 1164 (35.6)<br>No: 2103 (64.4)<br>*96.4% complete* |
| Concurrent MTX, n (%) | Yes: 135 (7.4)<br>No: 1694 (92.6)<br>*97.8% complete* | Yes: 31 (4.0)<br>No: 749 (96.0)<br>*95.7% complete* | Yes: 4 (4.7)<br>No: 82 (95.3)<br>*99.9% complete* | Yes: 21 (5.5)<br>No: 360 (94.5)<br>*97.9% complete* | Yes: 13 (6.4)<br>No: 191 (93.6)<br>*99.0% complete* | Yes: 60 (7.2)<br>No: 773 (92.8)<br>*99.0% complete* | Yes: 104 (7.1)<br>No: 1371 (92.9)<br>*98.4% complete* | Yes: 229 (7.0)<br>No: 3052 (93.0)<br>*96.9% complete* |
| Comorbidities, n (%) | Yes: 133 (8.2)<br>No: 1490 (91.8)<br>*86.7% complete* | Yes: 48 (7.4)<br>No: 598 (92.6)<br>*79.3% complete* | Yes: 6 (6.9)<br>No: 81 (93.1)<br>*100% complete* | Yes: 28 (8.4)<br>No: 305 (91.6)<br>*85.6% complete* | Yes: 12 (5.8)<br>No: 194 (94.2)<br>*100% complete* | Yes: 80 (9.8)<br>No: 740 (91.2)<br>*97.5% complete* | Yes: 87 (6.4)<br>No: 1273 (93.6)<br>*90.7% complete* | Yes: 239 (8.1)<br>No: 2694 (91.9)<br>*86.6% complete* |
| Mean baseline PASI, no. (SD) | 9.8 (7.3)<br>*73.4% complete* | 9.1 (7.7)<br>*69.0% complete* | 9.7 (7.7)<br>*83.9% complete* | 10.4 (7.6)<br>*77.9% complete* | 10.0 (7.7)<br>*80.6% complete* | 10.3 (7.2)<br>*69.7% complete* | 10.6 (7.4)<br>*79.1% complete* | 9.9 (7.2)<br>*70.0% complete* |
| Mean baseline DLQI, no. (SD) | 11.7 (7.1)<br>*65.9% complete* | 9.9 (7.4)<br>*54.1% complete* | 13.8 (6.3)<br>*52.9% complete* | 11.3 (7.3)<br>*51.9% complete* | 13.4 (6.7)<br>*60.2% completed* | 12.3 (7.2)<br>*70.7% complete* | 12.4 (7.3)<br>*68.3% complete* | 11.7 (7.2)<br>*66.8% complete* |
| Bio-Naïve, n (%) | 1298/2036 (63.7)<br>*100% complete* | 632/921 (68.6)<br>*100% complete* | 3/87 (3.4)<br>*100% complete* | 199/435 (45.7)<br>*100% complete* | 27/209 (12.9)<br>*100% complete* | 390/851 (45.8)<br>*100% complete* | 838/1632 (51.3)<br>*100% complete* | ----- |

There were no drastic differences in basic patient characteristics across different biologic treatments, with an exception of concurrent diagnosis of PsA, which was the lowest for patients treated with ustekinumab (23.5%) and the highest in the infliximab group (48.7%). Another difference was the proportion of treatment series administered to bio-naive patients (i.e., patients not previously exposed to a biologic), which varied from 3.4% in the guselkumab group to 68.6% for etanercept.

The majority (60.1%) of all registered treatment series were discontinued. The main reason for drug discontinuation was lack of efficacy (57.7%), followed by “other”, (37.2%) which comprised patients who were lost to follow up, changed the site of treatment, or for whom the reason was not entered to the database. Discontinuation due to adverse events was very low (1.6%); patient death was 1.1% and disease remission was 2.4%.

### Biologic Survival

To adjust for the confounders, we performed Cox regression analysis with the hazard ratios, using drug discontinuation as the outcome and adalimumab as reference **(Figure 1, Table S1)**. Comparing to adalimumab as a benchmark, the biologics associated with the lowest risk of discontinuation were ustekinumab (HR 0.61, 95% CI 0.52-0.72, P < 0.001) and ixekizumab (HR 0.61, 95% CI 0.39-0.97, P = 0.035), while etanercept was found to have the highest risk of biologic discontinuation (HR 1.57, 95% CI 1.34-1.88, P < 0.001). In addition, previous biologic therapy and the sex were also highly significant; there was significantly higher drug survival in bio-naïve males. Weight and baseline PASI score were statistically significant predictors, but their contribution was negligible in comparison with the three major predictors of type of drug, sex, and previous therapy **(Figure 1)**.

**Figure 1.**
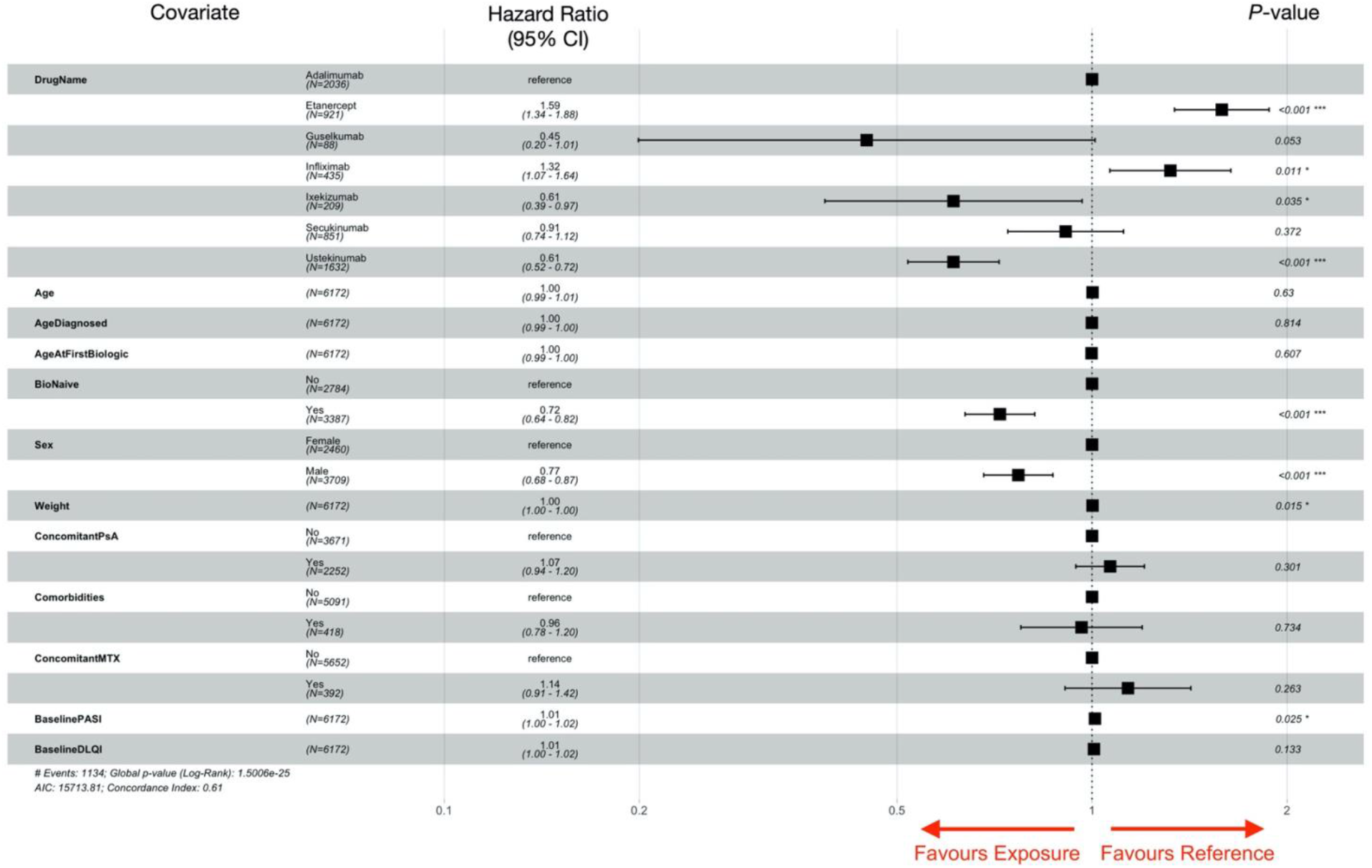
Forest plot of adjusted hazard ratios (HR) for risk of biologic discontinuation. For categorical variables, a HR greater than 1 indicates an increased risk of discontinuation, and a HR less than 1 indicates a decreased risk of discontinuation. For continuous variables, a HR greater than 1 indicates an increased risk of discontinuation with increases in the value of the variable, and a HR less than 1 indicates a decreased risk of discontinuation with increases in the value of the variable. A *P*-value of < 0.05 was considered statistically significant.

To provide the clinician with a practical tool to assess the individual risk of discontinuation, we constructed a medical nomogram comprising the clinically relevant variables identified by Cox regression. Using these salient predictors, it is possible to estimate a patient’s probability of 1-year, 3-year, and 5-year survival on a certain biologic given their characteristics **(Figure 2)**. To assess the performance of this tool, we calculated the area under the ROC curve (AUC) which is an aggregate measure of performance across all possible classification thresholds. AUC of 0.5 suggests no discrimination, the values between 0.7 to 0.8 are considered acceptable, whereas AUC >0.8 are considered excellent.^23^ The AUC based on the Cox regression is 0.61, which indicates low discriminatory value, but better than a random guess.

**Figure 2.**
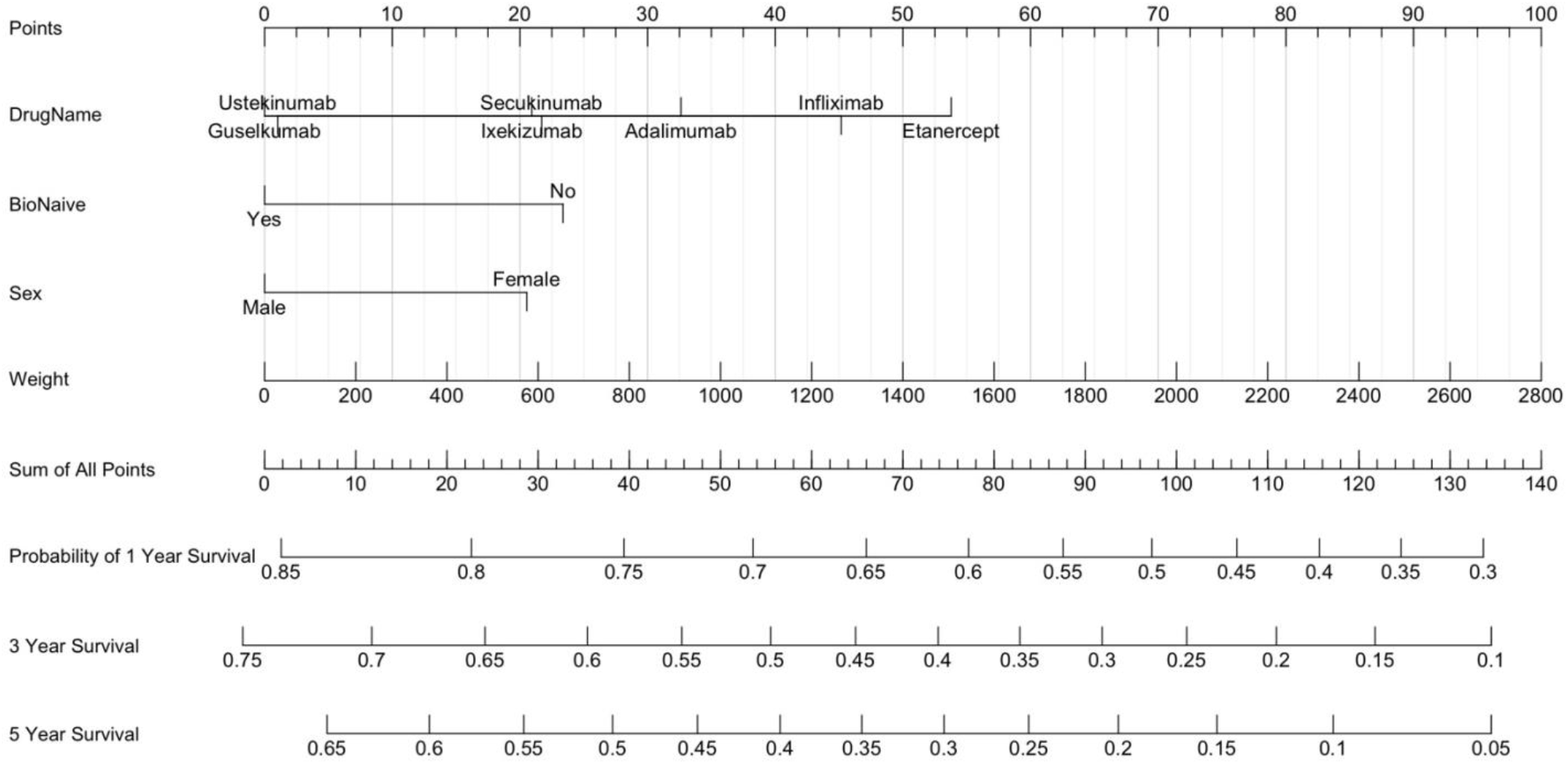
Nomogram for predicting probability of 1-year, 3-year, and 5-year biologic drug survival. To use the nomogram, draw a vertical line upwards from each variable to the ‘Points’ axis at the top of the figure. Record the number of points that each variable accrues, and manually sum the total points of all the variables. To estimate the survival probability at each aforementioned timepoint, proceed to the ‘Sum of All Points’ axis. Draw a vertical line downwards from the number calculated when adding all of the points together. This will provide the probability of 1-year, 3-year, and 5-year survival of a certain biologic based on a patient’s specific characteristics.

### Machine Learning Accurately Predicts the Risk of Biologic Discontinuation

We applied six different ML algorithms to test their accuracy in predicting the risk of drug discontinuation. As shown in **Table 2**, all algorithms predicted the 5-year likelihood of discontinuation with high accuracy ranging from 65.3% for Naive Bayes to 77.5% for Random Forest and Gradient Boosted Tree, respectively. Thus, the most efficient ML algorithms were able to predict the treatment outcomes with less than 23% classification error, only utilizing basic patient information routinely available to every clinician **(Figure 3, Table S2-S3)**. Similar to Cox regression analysis, the most important predictive parameters were: drug type (weight 0.247), sex (weight 0.076), and the body weight (weight 0.069), however, all clinical data contributed to the predictive model. The AUC measured for the Gradient Boosted Tree was 0.85, which is an indicator of excellent performance of the algorithm.^23^

**Table 2.** Machine learning algorithms with accuracy and training time.

| Model | Accuracy | Total Time | Training Time |
| --- | --- | --- | --- |
| Naïve Bayes | 65.3% | 104 s | 11 ms |
| Generalized Linear Model | 73.2% | 65 s | 23 ms |
| Deep Learning | 70.3% | 124 s | 691 ms |
| Decision Tree | 74.9% | 57 s | 7 ms |
| Random Forest | 76.1% | 787 s | 121 ms |
| Gradient Boosted Trees | 77.5% | 118 s | 135 ms |

**Figure 3.**
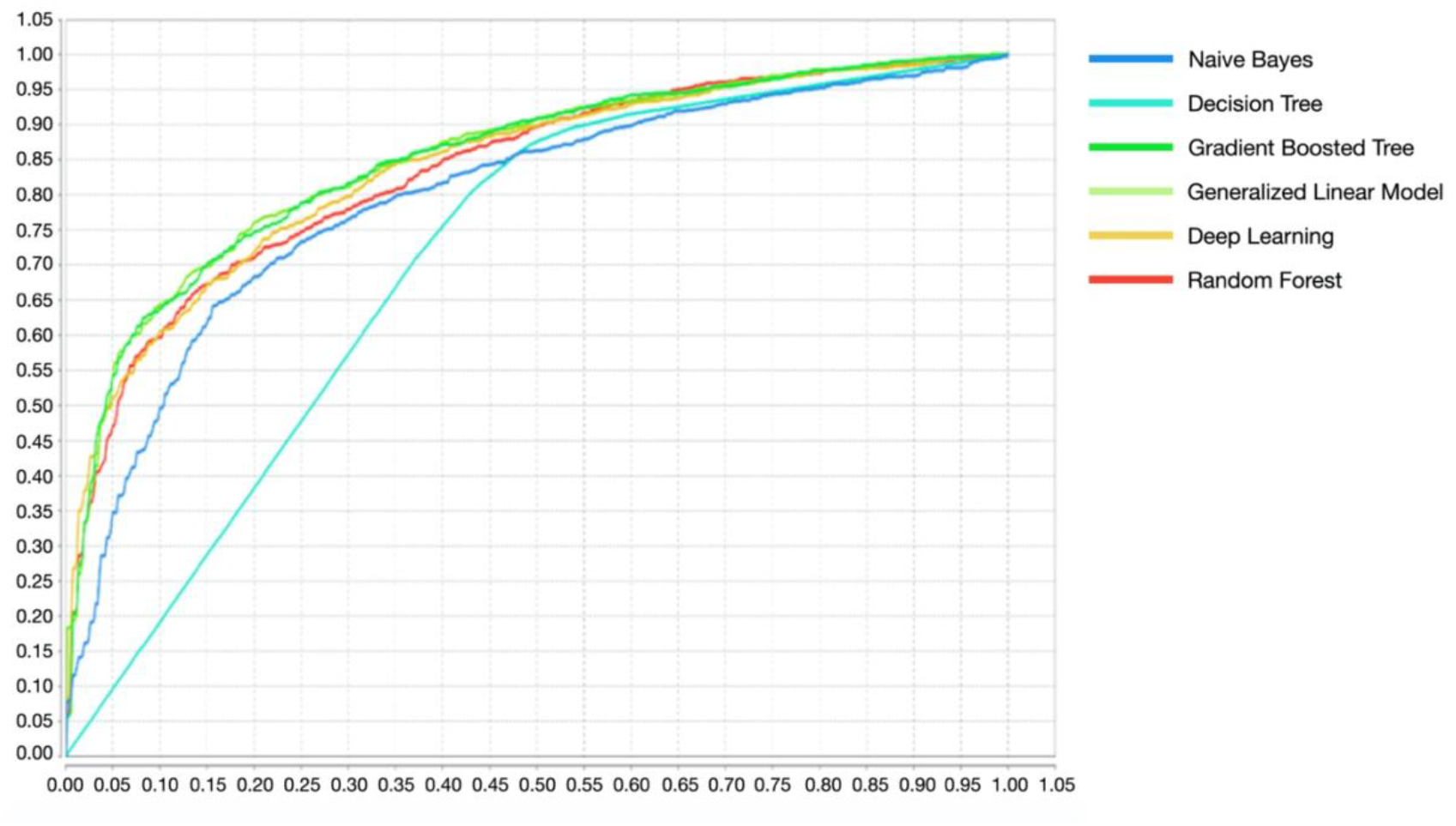
Receiver operating characteristic (ROC) curves reflecting the ability to predict the 5-year risk of discontinuation. Curves indicate performance of different machine learning (ML) models, as indicated in the legend.

## Discussion

Loss of the efficacy of biologics has been well documented in real-world data and is often of concern to the physician and the patient. Currently, dermatologists initiating biologic therapy for psoriasis lack personalized medicine tools to predict individual outcomes. Accurate prediction of the long-term efficacy of biologics represents an unmet need in clinical dermatology.

In our study, we assessed two potentially applicable predictive tools based on statistical modeling (Cox regression) and on ML. We found that even the most basic clinical information (sex, weight, drug name, previous exposure to a biologic) allowed us to predict the risk of discontinuation using a simple nomogram **(Figure 2)**. However, the discriminatory power, measured using AUC, was low to moderate (0.61), highlighting large variation in predicting the outcomes with statistical modeling.

ML explores non-linear, high-dimensional relationships between different features and has already shown considerable promise in the personalized assessment of therapeutic outcomes. The computation of risk of drug discontinuation using ML approach outperformed the Cox regression-based approach, although the utility of different algorithms varied considerably.

A limitation of this study was the observational design of the Dermbio registry. Validation of our ML algorithm should include data from other registries because treatment patterns and reasons of discontinuation cannot be standardized easily for different countries. For example, biologic dose increase is an effective method for optimizing the response,^24,25^ but might not be allowed in all countries. Patient weight was only a borderline significant risk factor, and this might be because dose escalations are allowed in clinical practice in Denmark, and the increased dose compensates for higher body mass. Also, in areas with difficult access to biologics, the threshold to discontinue an only partially efficient drug might be higher. Presence of available competing biologics may also influence therapeutic decisions.^26^ In spite of regional differences, analysis of drug survival rates yielded surprisingly high homogeneity regarding the 5-year discontinuation rate, risk factors for discontinuation, and the ranking of biologics with respect to long-term efficacy.^6,11^ Another limitation is the lack of validation of the ML algorithm in a prospective trial design. Even with our careful approach, incorporating a 5-fold cross validation technique, there is risk of model overfitting. However, this data may stimulate other clinical centers to develop their own ML tools to predict clinical outcomes, taking the regional differences into consideration. Finally, it is conceivable that a richer dataset would improve ML performance. The sophisticated ML systems developed for risk stratification rely on multiple variables. For example, the ML-based algorithm for prediction of adverse events following an acute coronary syndrome relies on 25 clinical parameters, whereas our ML engine utilizes only 10 basic clinical features extractable from Dermbio.

In conclusion, we have developed two predictive models assessing the chance of biologic drug discontinuation – a model based on statistical impact of risk factors and a ML-based tool. We show that the ML approach is superior and is a potentially applicable personalized medicine tool assisting dermatologists in optimal drug selection and patient communication.

## Data Availability

All data produced in the present study are available upon reasonable request to the authors.

