## Supplementary material for "Machine Learning Model for Predicting Outcomes of Biologic Therapy in Psoriasis": Table S1, Table S2, Table S3

**Table S1.** Multivariate Cox proportional hazards analysis output.

| Covariances | Coefficient | *P*-value | HR | 95% CI | |
|---|---|---|---|---|---|
| | | | | Lower | Upper |
| Biologic | | | | | |
| Adalimumab | Reference | Reference | Reference | Reference | Reference |
| Etanercept | 0.461 | < 0.001 | 1.586 | 1.341 | 1.876 |
| Guselkumab | -0.801 | 0.053 | 0.449 | 0.199 | 1.011 |
| Infliximab | 0.278 | 0.011 | 1.321 | 1.065 | 1.638 |
| Ixekizumab | -0.493 | 0.035 | 0.634 | 0.387 | 0.965 |
| Secukinumab | -0.094 | 0.372 | 0.911 | 0.741 | 1.118 |
| Ustekinumab | -0.493 | < 0.001 | 0.611 | 0.520 | 0.719 |
| Age | 0.002 | 0.630 | 1.002 | 0.995 | 1.010 |
| Age at Psoriasis Diagnosis | -0.001 | 0.814 | 0.999 | 0.995 | 1.004 |
| Age at First Biologic | -0.002 | 0.607 | 0.998 | 0.992 | 1.005 |
| Bio-Naïve | -0.328 | < 0.001 | 0.721 | 0.637 | 0.816 |
| Sex (Male) | -0.262 | < 0.001 | 0.769 | 0.680 | 0.870 |
| Weight (kg) | 0.002 | 0.015 | 1.002 | 1.000 | 1.003 |
| Concomitant PsA | 0.064 | 0.301 | 1.066 | 0.944 | 1.205 |
| Comorbidities | -0.037 | 0.734 | 1.038 | 0.777 | 1.195 |
| Concomitant MTX | 0.128 | 0.263 | 1.136 | 0.909 | 1.421 |
| Baseline PASI | 0.010 | 0.025 | 1.010 | 1.001 | 1.019 |
| Baseline DLQI | 0.007 | 0.134 | 1.007 | 0.998 | 1.016 |

HR, hazard ratio; CI, confidence interval; PsA, psoriatic arthritis; MTX, methotrexate; PASI, Psoriasis Area and Severity Index; DLQI, Dermatology Life Quality Index

**Table S2.** Detailed results for gradient boosted trees.

| Criterion | Value | Standard Deviation |
|---|---|---|
| Accuracy | 77.5% | ±1.0% |
| Classification Error | 22.5% | ±1.0% |
| AUC | 85.1% | ±0.6% |
| Precision | 84.5% | ±1.3% |
| Recall | 80.7% | ±2.4% |
| F Measure | 82.5% | ±1.1% |
| Sensitivity | 80.7% | ±2.4% |
| Specificity | 71.5% | ±1.8% |

**Table S3.** Confusion matrix for gradient boosted trees.

| | True ‘continue’ | True ‘stopped | Class ‘precision’ |
|---|---|---|---|
| Predicted ‘continue’ | 582 | 302 | 65.84% |
| Predicted ‘stopped’ | 232 | 1262 | 84.47% |
| Class ‘recall’ | 71.50% | 80.69% | ----- |
